# Rapid, accurate, nucleobase detection using FnCas9

**DOI:** 10.1101/2020.09.13.20193581

**Authors:** Mohd. Azhar, Rhythm Phutela, Manoj Kumar, Asgar Hussain Ansari, Riya Rauthan, Sneha Gulati, Namrata Sharma, Dipanjali Sinha, Saumya Sharma, Sunaina Singh, Sundaram Acharya, Deepanjan Paul, Poorti Kathpalia, Meghali Aich, Paras Sehgal, Gyan Ranjan, Rahul C. Bhoyar, Indian CoV2 Genomics & Genetic Epidemiology (IndiCovGEN) Consortium, Khushboo Singhal, Harsha Lad, Pradeep Kumar Patra, Govind Makharia, Giriraj Ratan Chandak, Bala Pesala, Debojyoti Chakraborty, Souvik Maiti

**Author notes:** These authors contributed equally (Mohd. Azhar, Rhythm Phutela, Manoj Kumar, Asgar Hussain Ansari, Debojyoti Chakraborty, Souvik Maiti). Correspondence (D.C.); (S.M.).

## Abstract

Rapid detection of pathogenic sequences or variants in DNA and RNA through a point-of-care diagnostic approach is valuable for accelerated clinical prognosis as has been witnessed during the recent COVID-19 outbreak. Traditional methods relying on qPCR or sequencing are difficult to implement in settings with limited resources necessitating the development of accurate alternative testing strategies that perform robustly. Here, we present FnCas9 Editor Linked Uniform Detection Assay (FELUDA) that employs a direct Cas9 based enzymatic readout for detecting nucleotide sequences and identifying nucleobase identity without the requirement of trans-cleavage activity of reporter molecules. We demonstrate that FELUDA is 100% accurate in detecting single nucleotide variants (SNVs) including heterozygous carriers of a mutation and present a simple design strategy in the form of a web-tool, JATAYU, for its implementation. FELUDA is semi quantitative, can be adapted to multiple signal detection platforms and can be quickly designed and deployed for versatile applications such as infectious disease outbreaks like COVID-19. Using a lateral flow readout within 1h, FELUDA shows 100% sensitivity and 97% specificity across all range of viral loads in clinical samples. In combination with RT-RPA and a smartphone application True Outcome Predicted via Strip Evaluation (TOPSE), we present a prototype for FELUDA for CoV-2 detection at home.

**Single sentence summary:** A method to identify nucleotide sequence or nucleobase identity using FnCas9 and its implementation in the rapid and accurate diagnosis of SARS-CoV-2

## INTRODUCTION

The rise of CRISPR Cas9 based approaches for biosensing nucleic acids has opened up a broad diagnostic portfolio for CRISPR products beyond their standard genome editing abilities^1,2^. In recent times, CRISPR components have been successfully used to detect a wide variety of nucleic acid targets such as those obtained from pathogenic microorganisms or disease-causing mutations from various biological specimens^3-10^. At the heart of such a detection procedure lies the property of CRISPR proteins to accurately bind to target DNA or RNA, undergo conformational changes leading to cleavage of targets generating a reporter-based signal outcome^11-15^. To enable such a detection mechanism to be safe, sensitive and reproducible across a large variety of targets, the accuracy of DNA interrogation and subsequent enzyme activity is extremely critical, particularly when clinical decisions are to be made based on these results^1,2^.

Current technologies relying on using CRISPR components for nucleic acid detection can sense the identity of the target either through substrate cleavage mediated by an active CRISPR ribonucleoprotein (RNP) complex or by binding through a catalytically inactive RNP complex. Cleavage outcomes are then converted to a reporter-based readout with or without signal amplification^1^. Among the CRISPR proteins that have been used so far, Cas12 and Cas13 have obtained Emergency Usage Authorization (EUA) for diagnostic use during the COVID-19 pandemic^16-17^. However each of these approaches has its own strengths and limitations that are related to sensitivity to mismatches and ease of design for wide variety of targets. For example, to genotype individuals with high confidence, including careers of a single nucleotide variant (SNV), Cas12 requires individual sgRNA design and optimization for every target^7^ which increases the complexity and thus the time taken for design and deployment of a diagnostic test. Importantly, both Cas12/Cas13 unleash a secondary reporter activity upon activation, leading to possible loss of information about starting copy numbers of the target^16-17^. Taken together, development of a detection pipeline based on a highly specific Cas protein with a direct binding or cleavage based readout can significantly increase the sensitivity of detection and reduce the time and cost of CRISPR based diagnostics (CRISPRDx), This is especially crucial for point-of-care (POC) applications where complex experimentation or setup of reaction components are not feasible.

We have recently reported a Cas9 ortholog from *Francisella novicida* (FnCas9) showing very high mismatch sensitivity both under *in vitro* and *in vivo* conditions^18-20^. This is based on its negligible binding affinity to substrates that harbor mismatches, a property that is distinct from engineered Cas proteins showing similar high specificity^21^. We reasoned that FnCas9 mediated DNA interrogation and subsequent cleavage can both be adapted for accurately identifying any single nucleotide variants (SNVs) provided that the fundamental mechanism of discrimination is consistent across all sequences. We name this approach FnCas9 Editor Linked Uniform Detection Assay (FELUDA) and demonstrate its utility in various pathological conditions including genetic disorders. Notably, due to its ease of design and implementation for new targets, we were successful in deploying it for a ready-to-use diagnostic kit during the COVID-19 outbreak that has successfully completed regulatory validation in India^22-24^.

## RESULTS

### FELUDA distinguishes between nucleotide sequences differing by 1 mismatch

To identify a SNV with high accuracy, we first sought to investigate if FnCas9 can be directed to cleave the wild type (WT) allele at a SNV by placing an additional MM in the sgRNA sequence specific to the SNV (Supplementary Figure 1A). To test this, we selected sickle cell anemia (SCA), a global autosomal recessive genetic disorder caused by a point mutation (GAG>GTG) ^25-26^ By fixing the position of the SNV and walking along the entire length of the sgRNA, we discovered that two mismatches at the PAM proximal 2^nd^ and 6^th^ positions completely abrogated the cleavage of the SNV target while leaving the WT target intact (Supplementary Figure 1B). We tested FELUDA on DNA cloned from 6 SCA patients and a healthy control and obtained a clearly identifiable signature for SNV in every case (Supplementary Figure 1C). Importantly, the same design principle for searching and discriminating can be universally applied to other Mendelian SNVs without the need for optimization (Supplementary Figure 1D). To aid users for quick design and implement FELUDA for a target SNV, we have developed a web tool JATAYU (Junction for Analysis and Target Design for Your FELUDA assay) that incorporates the above features and generates primer sequences for amplicon and sgRNA synthesis (https://jatayu.igib.res.in, Supplementary Figure 1E).

**Fig. 1:**
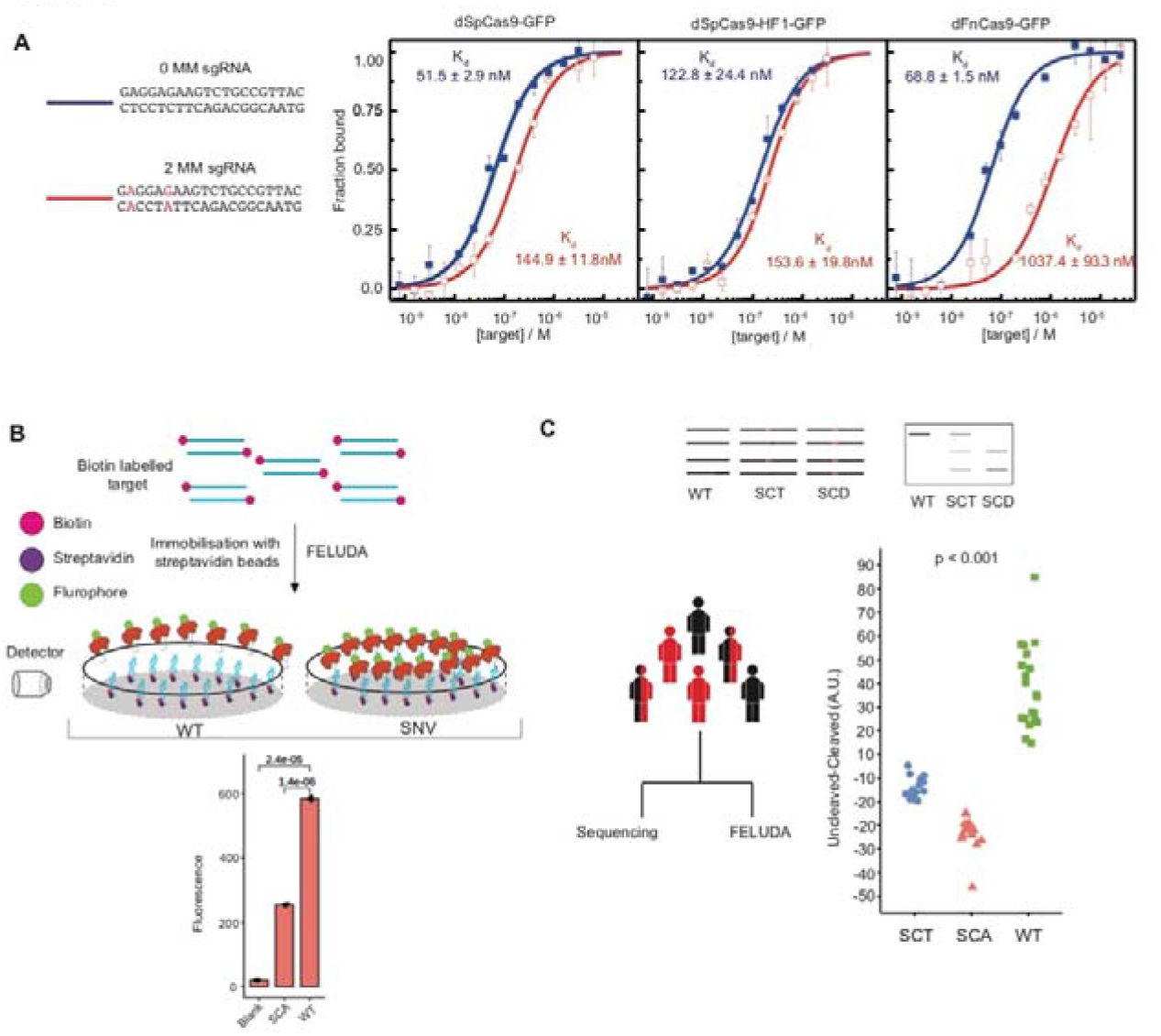
Schematic for FELUDA detection. A. Binding affinity experiments using Microscale Thermophoresis showing interaction of dSpCas9-GFP, dSpCas9-HF1-GFP and dFnCas9-GFP with substrates with 1 (blue) or 2 (green) mismatch (MM). Values are expressed as fraction bound protein (y-axis) with respect to varying concentrations of purified DNA substrate (Molar units, M, x-axis). Error bars represent SEM (2 independent experiments). **B**, Schematic for fluorescence based detection of sickle cell anemia mutation (SCA) using FELUDA. Error bars represent SD. Student’s t-test p values are shown (n= 3 independent measurements). **C**, Upper panel shows schematic of FELUDA for identifying carriers of SCA mutation. Lower panel shows blinded FELUDA results in a mixed cohort of individuals (n=49, one way ANOVA p value is shown). SCA, sickle cell anemia patient; SCT, sickle cell trait individuals; WT, normal subjects; SNV, single nucleotide variation; SEM, standard error of the mean.

### FELUDA diagnosis is adaptable to direct binding-based outcomes

We next sought to adapt FELUDA for fluorescent detection. Unlike Cas12/Cas13 based CRISPRDx platforms, FnCas9 is not reported to produce collateral activity on substrates, so FELUDA is not suitable for trans-cleavage signal output. To circumvent this, we envisioned FELUDA as a direct, non-cleavage, affinity-based method of detection which works with single nucleotide mismatch sensitivity. We first investigated if a catalytically inactive dead FnCas9 (dFnCas9) tagged with a fluorophore (GFP) is adept in sensing a point mutation at DNA using Microscale Thermophoresis (MST) (Figure 1A). We observed that using FELUDA specific sgRNAs (having 2 mismatches with WT), the WT substrate exhibited negligible dFnCas9-GFP binding (Kd = 1037.4 nM ± 93.3 nM) while the SCD substrate showed moderately strong binding (Kd = 187.2 nM ± 3.4 nM) (Figure 1A and Supplementary Figure 2A). This is consistent with the in vitro cleavage (IVC) outcomes on the two substrates (Supplementary Figure 1C). Importantly, both *Streptococcus pyogenes* Cas9 (SpCas9) and its engineered High-Fidelity variant (dSpCas9-HF1-GFP) showed strong binding to WT substrate with sgRNAs containing 2 mismatches (144.9 ± 11.8 nM and 153.6 ± 19.8 nM respectively) establishing that the inherent DNA interrogation properties of FnCas9 are responsible for discriminating single mismatched targets with very high specificity (Figure 1A). We then developed a pipeline to adapt FELUDA for an affinity-based fluorescent read-out system, where an amplification step generates biotinylated products that can then be immobilized on magnetic streptavidin beads (Figure 1B). Upon incubation with dFnCas9-GFP, enzymatic binding to the substrate leads to loss of fluorescence signal in the supernatant allowing FELUDA to discriminate between SCA and WT samples (Figure 1B).

### FELUDA accurately genotypes carriers of Mendelian variants

Although sickle cell trait (SCT) individuals are generally non-symptomatic, carrier screening is vital to prevent the spread of SCA in successive generations and is widely employed in SCA control programs in various parts of the world^26^. Since FELUDA outcomes are reflected by binding to substrate molecules, it resulted in clearly distinguishable signatures between the SCA, SCT and WT DNA obtained from patient’s saliva samples (Supplementary Figure 2B). To address the robustness of detecting the 3 genotype categories, we performed a blinded experiment using DNA obtained from 49 subjects with all three SCA genotypes from a CSIR-Sickle Cell Anaemia Mission Laboratory in Chhattisgarh state of India. Remarkably, FELUDA identified all three genotypes with 100% accuracy and the results perfectly matched with Sanger sequencing data generated on same samples in a different laboratory (CSIR-Center for Cellular and Molecular Biology, Genome Research on Complex Diseases Lab) (Figure 1C).

**Fig. 2:**
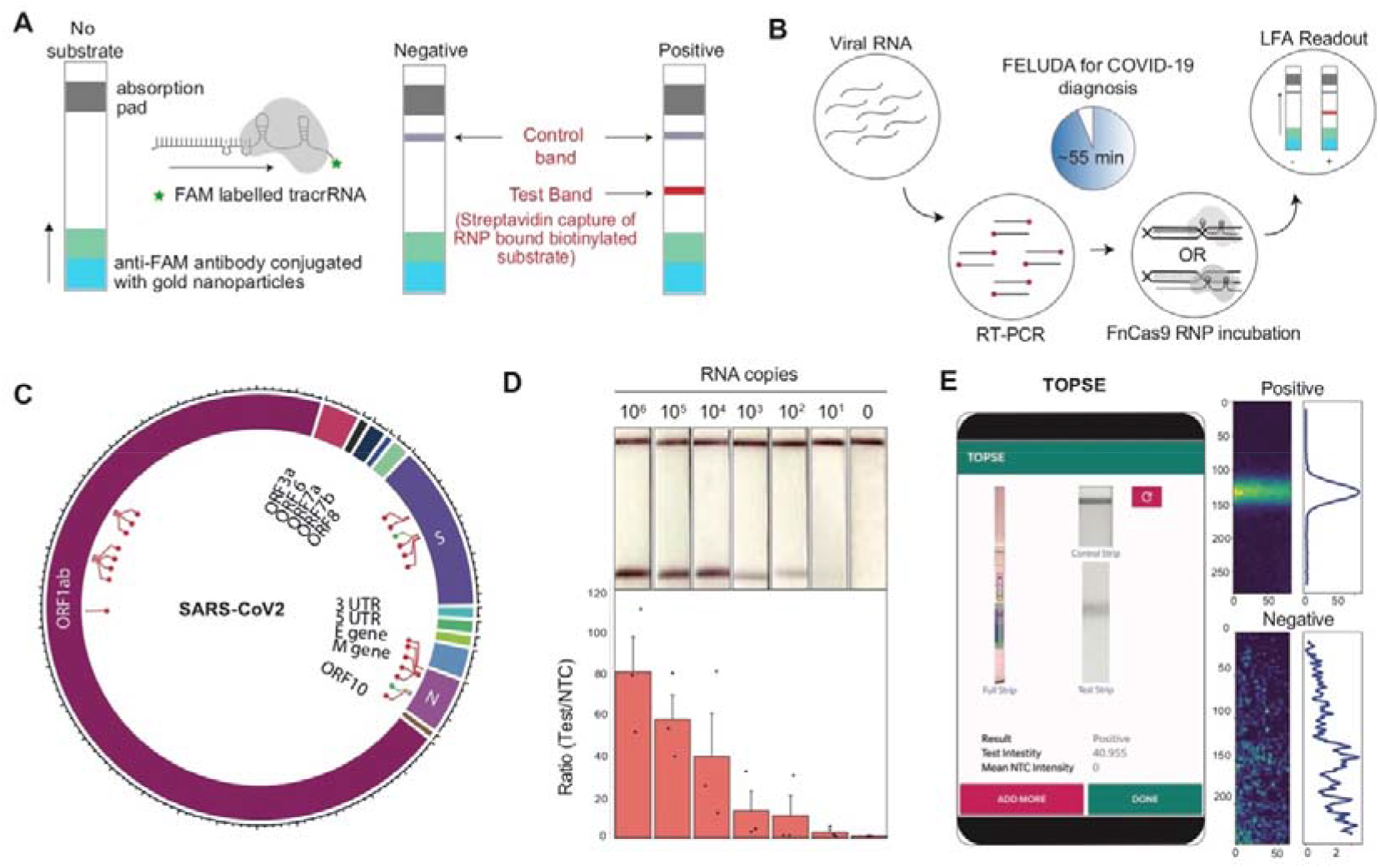
FELUDA for point-of-care nCOV-2 detection. A. Outline of lateral flow assay using FELUDA showing positions of control and test bands. **B**, Pipeline of FELUDA based detection for SARS-CoV-2 infection from patient samples. Individual steps are depicted. **C**, Plot showing the regions of SARS-CoV-2 RNA genome tested for FELUDA, successful regions represented in red. D, LOD of FELUDA in purified N gene target RNA. Top panel shows representative LFA readout on strips, bottom panel shows Fluorescent intensity ratios. Error bars s.e.m. (n=3 independent experiments). E, Smartphone GUI for TOPSE showing representative output from a strip image (left). Positive and NTC preprocessed images are shown on the right.

### PAMmer based amplification releases PAM dependency on FELUDA design

We made several improvements to FELUDA for expanding and simplifying its detection spectrum. To detect non-PAM proximal SNVs we installed an in-built PAM site in the amplification step of FELUDA (PAMmer) and successfully validated this approach using 2 SNVs (A2142G and A2143G) present in *Helicobacter pylori* 23s *rRNA* gene which confers variable clarithromycin resistance in patients with gastric ulcers^27^”^29^(Supplementary Figure 2C-D). Next, we tried to reduce PAM dependency on sgRNA design further by exploring if FELUDA could be performed with a single mismatch in the sgRNA. In line with previous reports, we found that FnCas9 shows negligible cleavage with sgRNAs containing mismatches at the PAM distal end and in particular, mismatch at PAM distal 16^th^ base showed complete absence of cleavage or binding (Supplementary Figure 3A-B). We confirmed this strategy by targeting the SNV rs713598 (G>C) in different individuals and successfully identified their genotypes ^30^ (Supplementary Figure 3C). We also show that FELUDA based detection can work robustly across a wide temperature range and up to 3 days post thawing of reaction components (at room temperature). Thus, field studies using FELUDA can be conducted in diverse climatic conditions and reaction components can be successfully used following cold chain transportation (Supplementary Figure 4A-B).

### FELUDA on paper strip can accurately predict COVID-19 outcomes

The recent outbreak of Coronavirus disease 19 (COVID-19) due to SARS-CoV-2 virus provided an opportunity to expand the scope of above-mentioned approach of FELUDA and make a difference in the ongoing public health emergency throughout the world. In addition to general social distancing, identification of infected individuals and, screening their contacts for possible quarantine measures is one of the major steps in reducing community transmission of this virus^31-33^. Although quantitative Real-Time (qRT) PCR is considered a gold standard test for detecting active COVID-19 cases, such tests are expensive, have long turn-around times and require a dedicated qRT-PCR machine, hence is of limited utility in handling an emergency of this scale. We sought to repurpose FELUDA as a lateral flow assay (LFA) for the detection of SARS-CoV-2 that is low-cost, does not need complex instrumentation, and is highly accurate in diagnosis. To enable such a diagnosis on commercially available paper strips we enabled the chemistry of capturing RNP-bound biotinylated substrate molecules on a distinct test line of the paper strip using FAM labeled chimeric gRNA (Figure 2A and Supplementary Figure 5A). Using an optimized single step Reverse Transcription-PCR protocol followed by FELUDA, we developed an assay that can detect SARS-CoV-2 sequences from RNA samples within an hour (Figure 2B and Supplementary Figure 5B, Supplementary Note 1). We tested up to 21 targets across the SARS-CoV-2 RNA genome and identified two regions (in the viral N and S genes) which are present at high copy numbers^34-35^ and reported negligible number of mutations in publicly available datasets^36-37^ (63,997, GISAID submissions till September 7, 2020) (Figure 2C, Supplementary Figure 6A. Supplementary Table 1). Through extensive optimization of PCR and reaction components, FELUDA reached a limit of detection (LOD) of ~10 copies of purified viral sequence (Figure 2D, Supplementary Figure 6B). Upon gradual dilution of patient RNA, both FELUDA and qRT-PCR was able to detect samples till the same dilution range (Supplementary Figure 6C). Since visual detection can occasionally have an operator-bias, particularly when the signal is very faint, we developed a smartphone app TOPSE (True Outcome Predicted via Strip Evaluation) to assist detection by returning a predictive score based on background correction (Figure 2E, Supplementary Note 2, and Supplementary Movie 1).

### FELUDA shows high concordance with gold standard qRT-PCR in detecting nCoV-2 infection

FELUDA detection is semi-quantitative (due to stoichiometric binding of FnCas9 RNP:target) and therefore shows a strong negative correlation between Ct values and signal intensities (Figure 3A). This makes it uniquely placed among CRISPRDx platforms to accurately predict the viral load in patient samples with high reproducibility between assays (Figure 3B)^16-17^.

**Fig. 3:**
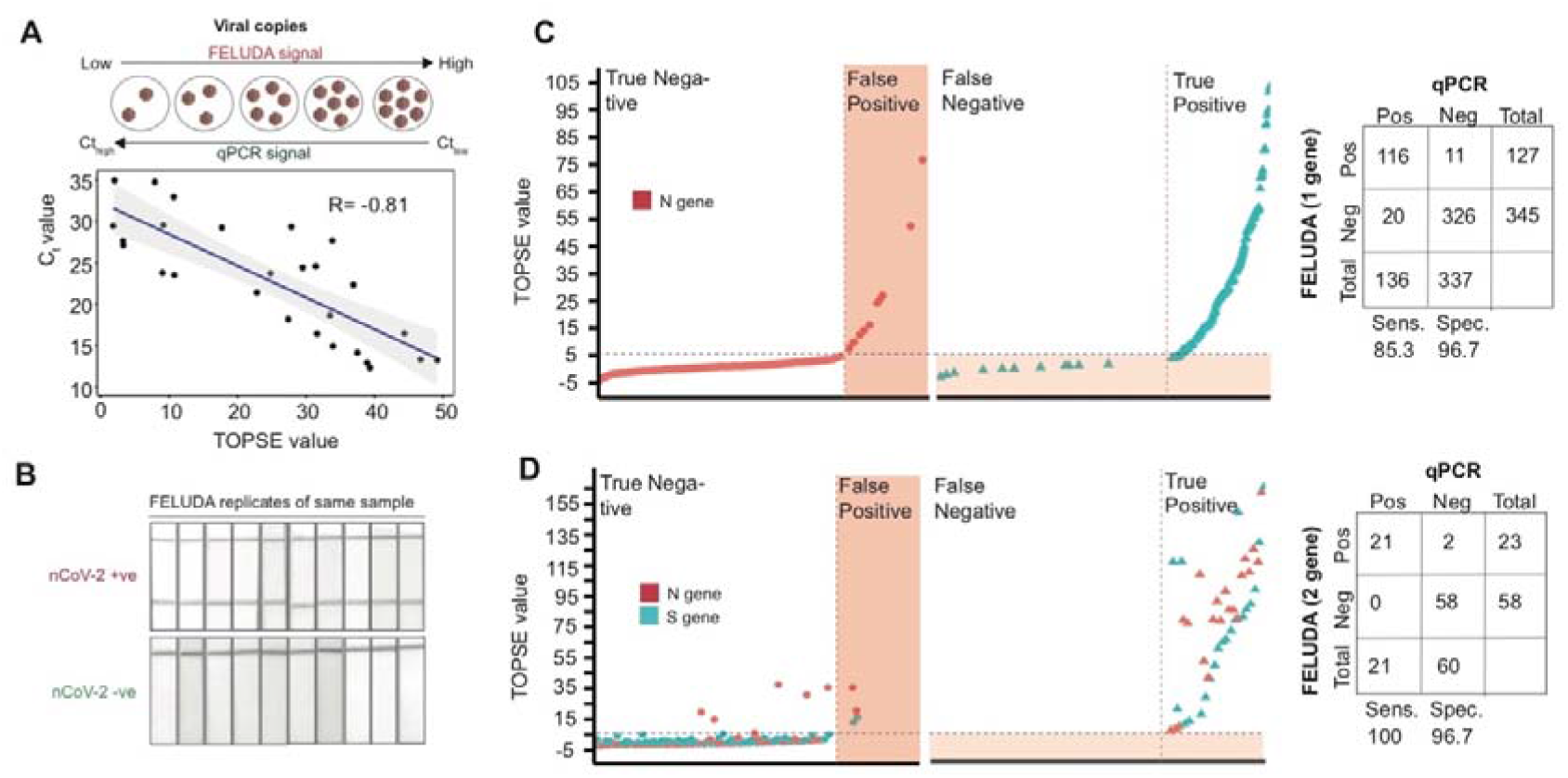
FELUDA shows high concordance with qRT-PCR for SARS-CoV-2 detection A. FELUDA readouts are semiquantitative. Correlation between Ct values (E gene) and TOPSE values are shown (n=27). **B**, Strong reproducibility between repeated FELUDA runs on the same positive or negative sample is shown. **C**, One gene (N gene) FELUDA on clinical samples (x axis) showing distribution of TOPSE values (y axis). Analyzed results represented on the right. D, Two gene (N and S genes) FELUDA on clinical samples (x axis) showing distribution of TOPSE values (y axis). Analyzed results represented on the right.

We first performed FELUDA using one gene (N), single pass assay on qRT-PCR confirmed 473 samples and obtained a sensitivity of 85.3% (116/136) and specificity of 96.7% (326/337) with qRT-PCR (Figure 3C, Supplementary Dataset 1). Among the samples that showed discordance between FELUDA with qRT-PCR, 8 false negative FELUDA samples and 10 false positive FELUDA samples were picked up for a repeat evaluation by FELUDA and qRT-PCR. Surprisingly a repetition of FELUDA with double the amount of RNA yielded positive signals in 6/8 (75%) of samples and a repeat qPCR yielded positive signals in 5/10 (50%) of initially classified negative samples (Supplementary Figure 7A-B). This underscores the error rates seen in a single run assay and has been reported elsewhere^38^. To improve FELUDA accuracy, we combined assays for both N and S genes, doubled the starting RNA amount and performed FELUDA with 81 qRT-PCR confirmed samples. Remarkably we obtained a sensitivity of 100% and a specificity of 97% and were able to accurately detect samples up to a high Ct value of 37 showing the robustness of the assay (Figure 3D, Supplementary Figure 7C, Supplementary Dataset 2). Based on the above results, we have completed the non-exclusive licensing of FELUDA and the technology has successfully concluded third party evaluation and regulatory validation in India.

### FELUDA can be adapted to a point-of-care or home testing assay

Understanding the need for more testing, especially in the wake of rising numbers and predictions of a second wave of infection^39^, we have implemented a PCR machine free version of FELUDA using Recombinase Polymerase Amplification (RPA)^40,41^ that can detect SARS-CoV2 RNA in biological samples within 30 min (Figure 4A, Supplementary Note 1). Finally, to adapt FELUDA for possible home testing in the future, we have successfully developed a 15 min benchtop method for RNA extraction from saliva and an on-body 30 min RPA-FELUDA (tested using synthetic RNA fragments), thus generating an end-to-end instrumentation free testing protocol (Figure 4B-C, Supplementary Figure 7D, Supplementary Movie 2, Supplementary Note 1).

**Fig. 4:**
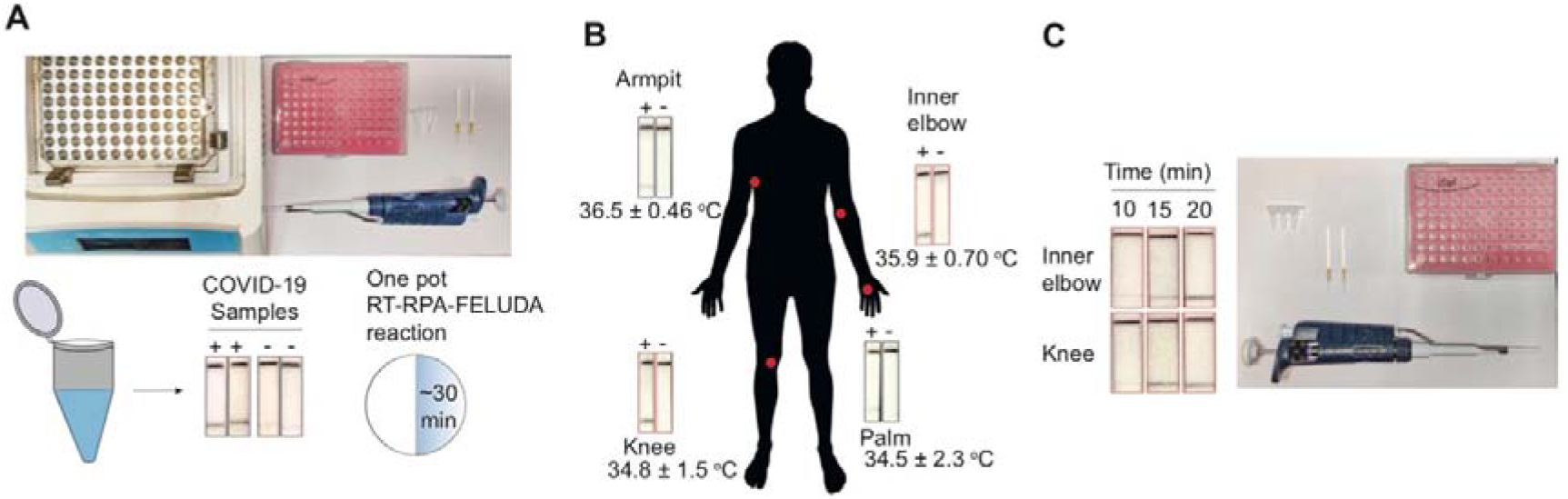
Prototype for FELUDA for possible home-testing A. One pot RT-RPA FELUDA. Top panel shows minimum requirements, bottom panel shows outcome for 2 representative samples. **B**, On-body RT-RPA-FELUDA, left figure shows variation of temperature in different zones of the body marked in red dots and corresponding RPA-FELUDA for a synthetic RNA fragment as starting material. **C**, Left panel shows representative FELUDA using on body RT-RPA from samples incubated in two different parts of the body. Right panel represents minimum requirements for FELUDA using on-body RT-RPA.

## DISCUSSION

In this study, we developed a FnCas9 based test for detecting a wide range of pathogenic SNVs and nucleotide sequences. We were able to design a scalable version of the test that is point-of-care, requires minimal equipment and validate the test on a large number of clinical samples. Our results suggest that FELUDA is suitable for SARS-CoV2 diagnosis with similar degree of accuracy as the currently accepted gold standard qRT-PCR test and can replace it in resource limited settings where qRT-PCR machine is not available or for mass scale community screening.

In this study we report three different modes of FELUDA readout. On the basis of cleavage, an agarose gel electrophoresis analysis gives the most accurate readouts at the level of single mismatch distinction, particularly in detecting carriers of SCA mutation. Although such a test is viable in lab settings and performed with 100% accuracy in blinded samples, this might not be suitable in places where conducting electrophoresis experiments is challenging. For such scenarios, a microfluidics-based fragment analyzer combined with a portable hand-held PCR might be more suitable^42^.

Genotyping carrier individuals with heterozygosity though sequencing-free methods systems is often complicated and requires extensive optimization of assay conditions^43^. The overall design of the assay is based on the high specificity of FnCas9 interaction with DNA substrates. We systematically analyzed mismatches at different positions of the sgRNA against a target and obtained a robust design parameter where presence of 2 mismatches (PAM proximal 2^nd^ and 6^th^) or 1 mismatch (PAM distal 16^th^) in the sgRNA abrogates the binding and cleavage of the target using FnCas9. This strategy has worked across all targets tested so far in this study suggesting that designing FELUDA sgRNAs follows a consistent principle which is independent of DNA target sequence. This has been implemented in the JATAYU tool. Designing and validation of complex targets (such as with repeats or variable GC content) need to be undertaken to establish this further.

There are certain limitations of FELUDA which the COVID-19 pandemic gave an opportunity to address. The LFA gives a semi-quantitative readout of viral load in a given sample. However, for accurate and sensitive diagnosis it is imperative that a large number of primer and sgRNA pairs are tested to account for robust readout, prevention of off-target amplification and minimizing chances of targeting mutated viral regions^44^. Of the 21 regions we targeted, only two (N and S gene) satisfied these criteria (Supplementary Table 1). Importantly, optimized conditions and high-quality PCR reagents are necessary to ensure robust and consistent FELUDA results (Supplementary Figure 6B).

We uncovered inconsistencies in qRT PCR Ct values between samples that were measured before and after freezing (Supplementary Figure 7A-B). This is relevant when comparing validation of new tests with results of qPCR-validated samples from the freezer, particularly those with high C_t_ values (Supplementary Figure 8). Indeed, our single gene FELUDA done on frozen samples and compared with qRT-PCR done at an earlier time point was less sensitive than double gene FELUDA on samples where simultaneous qPCR was done (Figure 3C-D).

The on-body RT-RPA FELUDA prototype combined with extraction-free RNA isolation can potentially bring the benefits of testing to home. Although RT-RPA is rapid and sensitive, it is also prone to aerosol-based contamination necessitating an efficient cold-chain transportation of pre-pipetted components to minimize user handling^45^. It is imperative to test this prototype on a larger number of clinical samples to establish its accuracy.

Taken together, FELUDA is an accurate and low-cost CRISPR based diagnostic assay to detect nucleic acids and variations. A two-gene FELUDA assay for SARS-CoV2 costs ~7 USD (Supplementary Table 2). Its single-mismatch sensitivity to nucleic acids expands its application portfolio to a large number of sectors not limited to healthcare. Its ease of design and implementation, as exemplified by its urgent deployment during the COVID-19 health crisis offers immense possibilities for rapid and wide-spread testing that has so far proven to be successful in spreading the progression of the disease in multiple countries.

## MATERIALS AND METHODS

### Study Design

The study was designed to evaluate the efficacy of FELUDA on clinical samples. The intent of the study was to develop a robust CRISPR diagnostic that can perform with high accuracy in point-of-care settings. For the SNV detection using FELUDA, SCD samples were collected after informed consent from volunteers in CSIR-Sickle Cell Anemia Mission Laboratory, Chhattisgarh Institute of Medical Sciences, Bilaspur 495001, Chhattisgarh, India and DNA was extracted at CSIR-Center for Cellular and Molecular Biology, Uppal Road, Hyderabad, Telengana 500007 where Sanger Sequencing was also done. Anonymized DNA samples were processed for FELUDA at CSIR-Institute of Genomics and Integrative Biology only after which results were validated between FELUDA and Sanger Sequencing. Anonymized DNA from gut biopsy samples to detect *Helicobacter Pylori* were collected after informed consent from volunteers in All India Institute of Medical Sciences, Ansari Nagar East, New Delhi 110029, India. Anonymized SARS CoV2 RNA samples were received from the diagnostic laboratory at CSIR-Institute of Genomics and Integrative Biology where clinical diagnosis was done prior to left-over sample being used in this study. For the one gene FELUDA assay, qRT-PCR results were retrieved from the diagnostic laboratory’s database. For the two gene assay, qRT-PCR was done simultaneously with FELUDA and reported in this study.

### Oligos

A list of all oligos used in the study can be found in Supplementary Table 3.

### Plasmid Construction

The sequences for the Hbb (WT and SCA) and GFP were PCR amplified and cloned into the TOPO-TA vector (Thermo Fisher Scientific) to be used as target sequence for Cas9 *in vitro* cleavage assays. 500bp sequences containing SNV for four Mendelian disorders Glanzman, Thrombasthenia, Hemophilia A (Factor VIII deficiency), Glycogen Storage Disease Type I and X-linked myotubular myopathy were ordered as synthetic DNA oligos (GenScript) and cloned into the pUC57 by EcoRV (NEB) to be used as target sequence for Cas9 *in vitro* cleavage assays. Similarly, a 500bp sequence flanking two SNVs (A2142G and A2143G) in *Helicobacter pylori* 23s *rRNA* gene were ordered as synthetic DNA oligos (GenScript) and cloned in pUC57 by EcoRV (NEB) to be used as target sequence for Cas9 *in vitro* cleavage assays.

### Site-directed mutagenesis to generate catalytically inactive FnCas9 (dFnCas9)

Catalytically inactive FnCas9 double mutants (D11A, H969A) were generated on pET28-His-10-Smt3-FnCas9 plasmid backbone (Acharya, S. et al. Proc. Natl. Acad. Sci. 116, 20959-20968 (2019)) by QuickChange II site directed mutagenesis kit (Agilent) following manufacturer’s protocol with some modifications.

### Protein purification

The proteins used in this study were purified as reported previously^18^. Plasmids containing FnCas9-WT, dFnCas9 and dFnCas9-GFP sequences were expressed in Escherichia coli Rosetta2 (DE3) (Novagen). The protein expressing Rosetta2 (DE3) cells were cultured at 37°C in LB medium (supplemented with 50mg/l Kanamycin) until OD600 reached 0.6 and protein expression was induced by addition of 0.5mM isopropyl β-D-thiogalactopyranoside (IPTG). The Rosetta2 (DE3) cells were further cultured at 18°C overnight and harvested by centrifugation. The E.coli cells were resuspended in lysis buffer (20mM HEPES, pH 7.5, 500mM NaCl, 5% glycerol supplemented with 1X PIC (Roche), 100ug/ml lysozyme and lysed by sonication and centrifuged. The lysate was affinity purified by Ni-NTA beads (Roche) and the eluted protein was further purified by size-exclusion chromatography on HiLoad Superdex 200 16/60 column (GE Healthcare) in 20 mM HEPES pH 7.5, 150 mM KCl, 10% glycerol, 1mM DTT. The concentration of purified proteins were measured by Pierce BCA protein assay kit (Thermo Fisher Scientific).The purified proteins were stored at -80°C until further use. Representative images of purified proteins used in the study are shown in Supplementary Figure 9.

### *In vitro* transcription (IVT)

*In vitro* transcription for sgRNAs/crRNAs were done using MegaScript T7 Transcription kit (ThermoFisher Scientific) following manufacturer’s protocol and purified by NucAway spin column (ThermoFisher Scientific). IVT sgRNAs/crRNAs were stored at -20°C until further use.

### Genomic DNA extraction

Human Genomic DNA was extracted from the blood using the Wizard Genomic DNA Purification kit (Promega) as per the manufacturer’s instructions.

Genomic DNA extraction from human saliva, 1ml of saliva was centrifuged at 13000 rpm followed by three washes with 1ml of 1X PBS. After washing, the pellet was lysed with 50^l of 0.2% Triton X100 at 95°C for 5 minutes. Then again centrifuged at 13000 rpm and supernatant was transferred into a fresh vial. A total volume of 1µl supernatant was used in PCR reaction or stored at -20 °C until further use.

Genomic DNA was extracted from the biopsy samples (15-20 mg) of patients infected with *Helicobacter Pylori* using DNeasy 96 PowerSoil Pro QIAcube HT Kit using Vortex Genei for the sample lysis.

### SCA FELUDA Detection assays

Sequences containing WT, SCT and SCA were amplified using primers with/without 5’ biotinylation from genomic DNA extracted from saliva or blood samples.

### Detection *via In vitro* Cleavage (IVC)

DNA or RNA converted to DNA were PCR amplified to be used as a substrate in *in vitro* cleavage assay, optimized in our previous study. ~100ng of purified DNA amplicon was incubated in reaction buffer (20 mM HEPES, pH7.5, 150mM KCl, 1mM DTT, 10% glycerol, 10mM MgCl_2_) along with reconstituted RNP complex (500nM) at 37 for 30 minutes and cleaved products were visualized on agarose gel.

### Detection *via* Fluorescence Detection

DNA regions from Hbb locus (WT & SCA) were amplified using biotinylated primers. dFnCas9-GFP: sgRNA (180 nM:540 nM) RNP complex was reconstituted at 25°C for 10 mins in reaction buffer (20 mM HEPES, pH7.5, 150mM KCl, 1mM DTT, 10% glycerol, 10mM MgCl2). Meanwhile, 6^L of the Dynabeads MyOne Streptavidin C1 (Thermo Fisher Scientific) were prepared. Beads were incubated with 1^M of biotinylated Hbb amplicon (WT & SCA) for 30 minutes in the reaction buffer. Further, dFnCas9-GFP RNP complex was incubated with the streptavidin bound PCR amplicon for 30 minutes. Emission spectra of unbound dFnCas9-GFP was measured using Monolith NT. 115 (NanoTemper Technologies GmbH, Munich, Germany) under 60% excitation power in blue filter (465-490nm excitation wavelength; 500-550nm emission wavelength) with medium MST power.

### Detection *via* Lateral Flow Detection

Chimeric gRNA (crRNA:TracrRNA) was prepared by mixing crRNA and synthetic 3’- FAM labelled TracrRNA in a equimolar ratio within annealing buffer (100mM NaCl, 50mM Tris-HCl pH 8 and 1mM MgCh), heated at 95LC for 2-5 minutes and then allowed to cool at room temperature for 15-20 min. Chimeric gRNA-dead FnCas9 RNP complex (500nM) was prepared by mixing them in buffer (20mM HEPES, pH7.5, 150mM KCl, 1mM DTT, 10% glycerol, 10mM MgCl_2_) and incubated for 10 min at RT. Target (WT & SCA) biotinylated amplicons were then incubated with the RNP complexes for 15 min at 37°C. Dipstick buffer was added to the reaction mix along with Milenia HybriDetect 1 lateral flow strip. Allow the solution to migrate into the strip for 2 minutes at room temperature and observe the result.

### FnCas9 cold chain activity

FnCas9-sgRNA complex (500nM) was prepared in a buffer (20mM HEPES, pH7.5, 150mM KCl, 1mM DTT, 10% glycerol, 10mM MgCl2) and incubated for 10 min at RT. Putting together RNP complexes along with DNA amplicons, IVC assays were performed at different temperatures ranging from 10°C to 50°C for 30 min. The reaction was then terminated using 1^l of Proteinase K (Ambion) and removing residual RNA by RNase A (Purelink), cleaved products were visualized on agarose gel.

### FnCas9 cold chain activity (upon storage at RT)

Post thaw FnCas9 for varied time points starting from 0Hrs to 100Hrs, FnCas9- sgRNA complex (500nM) was prepared in a buffer (20mM HEPES, pH7.5, 150mM KCl, 1mM DTT, 10% glycerol, 10mM MgCh). To monitor cleavage activity on linearized DNA plasmid, IVC assays were performed for 30 min with/without 10% sucrose. The reaction was then terminated using 1^l of Proteinase K (Ambion) and removing residual RNA by RNase A (Purelink), cleaved products were visualized on agarose gel.

### Microscale thermophoresis

For the binding experiment using Monolith NT. 115 (NanoTemper Technologies GmbH, Munich, Germany), dFnCas9-GFP protein along with 12% Urea-PAGE purified IVT sgRNAs were used. RNP complex (Protein:sgRNA molar ratio, 1:1) was prepared at 25^D^C for 10 min in buffer (20 mM HEPES, pH 7.5, 150mM KCl, 1mM DTT, 10mM MgCh). Target dsDNA were formed using HPLC purified 30bp ssDNA oligos (Sigma) by incubating them for 5 min at 95^D^C and then slow cool at 25^D^C. dsDNA target sequences varying in concentration (ranging from 0.7nM to 25|jM) were incubated with RNP complex at 37^D^C for 60 min in reaction buffer. NanoTemper standard treated capillaries were used for loading the sample. Measurements were performed at 25°C using 40% LED power in blue filter (465-490nm excitation wavelength; 500-550nm emission wavelength) and 40% MST power. Data was analysed using NanoTemper analysis software and plotted using OriginPro 8.5 software.Sanger Sequencing:

The sequencing reaction was carried out using Big dye Terminator v3.1 cycle sequencing kit (ABI, 4337454) in 10|jl volume (containing 0.5|jl purified DNA, 0.8|jl sequencing reaction mix, 2µl 5X dilution buffer and 0.6|jl forward/ reverse primer) with the following cycling conditions - 3 min at 95°C, 40 cycles of (10 sec at 95°C, 10 sec at 55°C, 4 min at 60°C) and 10 min at 4°C. Subsequently, the PCR product was purified by mixing with 12|jl of 125mM EDTA (pH 8.0) and incubating at RT for 5 min. 50|jl of absolute ethanol and 2|jl of 3M NaOAc (pH 4.8) were then added, incubated at RT for 10 min and centrifuged at 3800rpm for 30 min, followed by invert spin at <300rpm to discard the supernatant. The pellet was washed twice with 100|jl of 70% ethanol at 4000rpm for 15 min and supernatant was discarded by invert spin. The pellet was air dried, dissolved in 12|jl of Hi-Di formamide (Thermo fisher, 4311320), denatured at 95°C for 5 min followed by snapchill, and linked to ABI 3130xl sequencer. Base calling was carried out using sequencing analysis software (v5.3.1) (ABI, US) and sequence was analyzed using Chromas v2.6.5 (Technelysium, Australia).

### Disease selection from ClinVar Database

ClinVar dataset (version: 20180930) was used to find out disease variation spectrum that can be targeted by FELUDA^46^. SNVs which are situated 2bp upstream of the PAM sequence were extracted. Further, SNVs with valid OMIM ID having Pathogenic effects were filtered. Finally, variations with higher frequency in Indian Population were selected for the validation using customized python script.

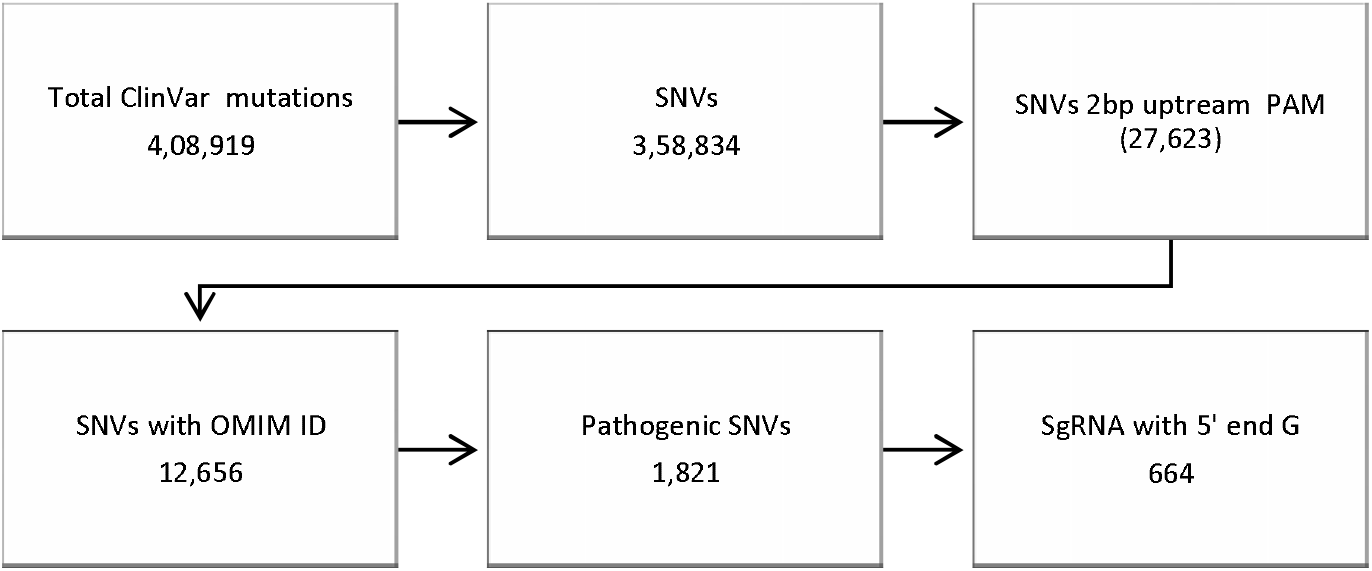

### JATAYU

JATAYU, web tool to design sgRNAs and primer for the SNV detection. When provided valid genomic DNA sequence with position and type of variation. JATAYU user interface has been created using Bootstrap 4 and jQuery. And works with a customized python-based Flask framework along with genome analysis tools like BWA (Burrows-Wheeler aligner) and bedtools^47-48^. Working script is available at https://github.com/asgarhussain/JATAYU.

### RT-PCR-FELUDA primer and crRNA design

To design, crRNA specific to SARS-COV 2, genome sequences were downloaded from GISAID (5). crRNAs were designed by searching any 20 nucleotides followed by NGG PAM. Further, to remove non-specific crRNA, off-target analysis was done by mapping them to human host viruses from Influenza Virus Database^37^ and human transcriptome (GENCODE GRCh38). Finally, 21 crRNAs with higher conservation frequency across SARS-COV 2, genome sequences in the GISAID dataset were selected^35^. Target PCR Primer sequences flanking these crRNAs are also investigated for off-targets on human transcriptome (GENCODE GRCh38).

Mutation frequency of the crRNA were routinely checked across GISAID datasets (63,997 genomes as of 7th Sep. 2020). Custom python script and SeqMap^49^ were used to design crRNAs and calling mutations (Supplementary Table 1).

### FELUDA Limit of detection (LOD)

Synthetic genomic Target for N gene was serially diluted (1:10, 7 times) from ~4×10^6^ copies/^l to perform FELUDA reaction. Test band intensity was calculated using TOPSE app (Repeated in three independent experiments).

### qRT-PCR SARS-CoV-2 detection

SARS-CoV-2 positive patient sample was titrated (1:10, 8 times), qRT-PCR was performed using STANDARD M nCoV Real-Time Detection kit (SD Biosensor) as per manufacturer’s protocol. Briefly, per reaction 3^l of RTase mix and 0.25^l of Internal Control A was added to 7^l of reaction solution. 5^l of each of the negative control, positive control, and patient sample nucleic acid extract was added to the PCR mixture dispensed in each reaction tube. The cycling conditions on instrument were as follows: Reverse transcription 50°C for 15 minute, Initial denaturation 95°C for 1 min, 5 Pre-amplification cycles of 95°C for 5 sec; 60°C for 40 sec followed by 40 amplification cycles of 95°C for 5 sec; 60°C for 40 sec. Fluorescence signals in FAM channel for qualitative detection of the new coronavirus (SARS-CoV-2) ORF1ab (RdRp) gene, JOE (VIC or HEX) channel for qualitative detection of the coronavirus E gene, and CY5 channel for detection internal reference.

### Extraction free detection of RNA from Saliva samples

Saliva RNA was extracted from 3 patient samples using RNeasy Kit (Qiagen). For extraction free detection, lysis buffer was prepared by adding 0.5% Triton X-100 in 50mM Tris Buffer pH 5. 0.2U/|jl RNase Inhibitor (EUROGENTEC) was added to this buffer solution. 200^l of saliva specimen was collected in a 1.5ml eppendorf tube. 100^l of prepared lysis buffer solution was pipetted to the tube and mixed by flicking. The tube was incubated at 95°C for 5 minutes on a dry bath after and left undisturbed on bench for 10 min.

One step qRT-PCR was performed by adding 1 ^l Reverse Transcriptase (Qiagen) to reaction mix with LightCycler® 480 SYBR Green I Master (Roche). *ACTB* specific primers at a final concentration of 0.2^M. qRT-PCR reactions were performed using 2ul of the kit extracted RNA and lysed supernatant for each patient. The cycling conditions on instrument were as follows: 1 cycle of Reverse transcription at 42°C for 15 minute, Initial denaturation 95°C for 5 min followed by 40 amplification cycles of 95°C for 10 sec; 60°C for 30 sec; 72 for 30.

## Data Availability

All data referred to in the manuscript can be obtained by mailing to

## Acknowledgments

We thank all members of Chakraborty and Maiti labs for helpful discussions and valuable insights. We are grateful to Mitali Mukerji, Rajesh Pandey and Mohd. Faruq (CSIR IGIB) for providing DNA samples used in the study. We are thankful to Pramod Kumar and Partha Rakhit (NCDC, New Delhi) for support during FELUDA optimization and the entire ADIUVO Diagnostics team (Chennai) for TOPSE development. We are grateful to Anurag Agrawal (CSIR IGIB) for critical insights during the study design. This study was funded by CSIR Sickle Cell Anemia Mission (HCP0008), TATA Steel CSR (SSP2001) and a Lady Tata Young Investigator award (GAP0198) to D.C.

## Contributions

D.C., S.M., M.A., R.P., M.K. and A.H.A conceived, designed and interpreted the experiments. A.H.A provided bioinformatics support. D.S. performed MST experiments. N.S. performed mismatch-based cleavage assays. R.P., M.K., R.R., S.G., S.S., S.Si, S.A., D.P and P.K. contributed to COVID testing. M.Ai. and K.S. contributed to studies for single mismatch discrimination using FELUDA. P.S., G.R., R.C.B, contributed to RNA isolation and qRT-PCR. H.L. and P.K.P. helped in the random screening of school children and identifying sickle cell anemia patients at Chhattisgarh. G.M. designed studies on detecting *H Pylori* variants. G.R.C performed sequencing of SCA patient samples. B.P. trained and designed TOPSE app. D.C. wrote the manuscript with inputs from S.M. and G.R.C.

## Ethics declarations

The present study was approved by the Ethics Committee, Institute of Genomics and Integrative Biology, New Delhi and Chhattisgarh Institute of Medical Sciences,Bilaspur. Two provisional patent applications have been filed in relation to this work. Mohd. Azhar is currently an employee of TATA Chemicals, India.

